# Cognitive enhancement: Effects of methylphenidate, modafinil and caffeine on latent memory and resting state functional connectivity in healthy adults

**DOI:** 10.1101/2021.11.07.21266019

**Authors:** Maxi Becker, Dimitris Repantis, Martin Dresler, Simone Kühn

## Abstract

Stimulants like methylphenidate, modafinil and caffeine have repeatedly shown to enhance cognitive processes such as attention and memory. However, brain-functional mechanisms underlying such cognitive enhancing effects of stimulants are still poorly characterized. Here, we utilized behavioral and resting-state fMRI data from a double-blind randomized placebo-controlled study of methylphenidate, modafinil and caffeine in 48 healthy male adults. The results show that performance in different memory tasks is enhanced, and functional connectivity (FC) specifically between the fronto-parietal (FPN) and default mode (DMN) network is modulated by the stimulants in comparison to placebo. Decreased negative connectivity between right prefrontal and medial parietal but also between medial temporal lobe and visual brain regions predicted stimulant-induced latent memory enhancement. We discuss dopamine’s role in attention and memory as well as its ability to modulate FC between large-scale neural networks (e.g. FPN and DMN) as a potential cognitive enhancement mechanism.

## Introduction

The umbrella term *cognitive enhancement* refers to interventions by which healthy individuals attempt to improve their cognitive functions, e.g. attention, cognitive control or memory (Repantis et al., 2010). Next to cognitive training, the use of psychoactive substances has become a popular way to obtain cognitive enhancement (Franke &Lieb, 2010; Talbot, 2009). Caffeine (CAF) in the form of caffeine-containing beverages is probably one of the most widely used prescription-free psychoactive substance worldwide (Ferré, 2008). But also prescription-stimulants such as methylphenidate (MPH, Ritalin®) and modafinil (MOD, Vigil®) are increasingly used by healthy individuals in the attempt to enhance cognition (Battleday & Brem, 2015; Compton et al., 2018). The mode of action as well as different cognitive enhancement effects of all three stimulants have been subject to several studies.

The natural stimulant CAF acts as a nonselective antagonist by blocking adenosine A_1_ and A_2_ receptors increasing energy metabolism (Koppelstaetter et al., 2010; Nehlig et al., 1992). Caffeine is most often used and shows strong effects as countermeasure to prolonged wakefulness (Walsh et al., 1990; for a review see Irwin et al., 2020). However, beneficial effects of CAF have been also reported on sustained attention, processing speed, vigilance and memory (Ullrich et al., 2015; Koppelstaetter et al., 2010; Nehlig, 2010; Borota et al., 2014; for a critical review on the cognitive effects of caffeine, see James & Keane, 2007; James, 2014).

MPH functions as a catecholamine reuptake inhibitor increasing extracellular dopamine in prefrontal, striatal and hippocampal regions and noradrenaline specifically in frontal brain regions by blocking their respective transporters due to binding to it (Berridge et la., 2006; Spencer et al., 2015; Kuczenski and Segal, 1997; Volkow et al., 2001). There is evidence that MPH enhances working memory and memory consolidation while evidence for processing speed, cognitive control and attention is rather mixed (Caviola& Faber, 2015; Linssen et al., 2014 or Repantis et al., 2010 for a review).

Similar to CAF, MOD is a wakefulness-promoting agent, meaning it is most often used to stave off the effects of sleep deprivation or excessive daytime sleepiness (Repantis et al., 2010; Bastoji & Jouvet, 1998; Boivin et al., 1993; Pigeau et al., 1995). Its precise mechanism is not entirely known up to date. Similar to MPH, MOD also elevates extracellular levels of catecholamines through inhibition of dopamine and noradrenaline transporters (Franke et al., 2017). However, MOD is believed to additionally affect other neurotransmitter systems such as promoting glutamate, serotonin, histamine pathways (Minzenberg & Carter 2008; Repantis et al., 2010). In general, MOD affects frontal lobe areas (Scoriels et al., 2013 for a review) and has been associated with improved attention, vigilance as well as memory and learning (Linssen et al., 2014; Repantis, 2010; Battleday & Brem, 2015, Müller et al., 2013; Randall et al., 2005).

Although the cognitive enhancement effect of all three stimulants is heavily dose-dependent (Woord et al., 2014), there seems to be converging evidence that single-dose intake can improve memory function in a healthy population (for different meta-analyses, see Linssen et al., 2014; Repantis, 2010; Ilieva et al., 2015). Despite the different primary modes of action of the stimulants, this poses the question whether there is a shared cognitive enhancement effect related to memory. One possible common neurobiological denominator could be that the stimulants directly or indirectly increase the extracellular levels of catecholamines in the brain. Although CAF does not exert its primary actions on the dopamine and noradrenaline systems, unlike MPH and MOD, it is assumed to indirectly modulate them via its antagonistic adenosine A_1_ receptors (Nehlig et al., 1992; Manalo & Medina, 2018; Volkow et al., 2015). For example, in vitro studies in animals have shown that CAF affects the local release of catecholamines, especially dopamine (Nehlig et al., 1992).

Although midbrain dopaminergic neurons mostly project to the basal ganglia and prefrontal cortex, evidence suggests that dopamine transmission plays an essential role in shaping large-scale neural networks underlying cognitive functions (Bentivoglio and Morelli, 2005; Rosenberg et al., 2016; Birn et al., 2019). For example, previous work found widespread effects of MPH, MOD and CAF on functional connectivity (FC). Single-dose MPH intake has been found to modulate functional network connectivity related to enhanced attention (Rosenberg et al., 2016) and modulate coupling between prefrontal regions with inhibitory and attention networks related to improved response inhibition (Pauls et al., 2012). Another study found that MPH reduced connectivity between default mode and dorsal attention, ventral attention, and visual networks in healthy adults (Sripada et al., 2013). MOD increased negative coupling between executive and the default mode network which was associated with improved cognitive control in alcohol-dependent patients (Schmaal et al., 2013). Single-dose MOD intake has also been found to increase FC between prefrontal and striatal areas (Cera et al. 2014). Finally, single-dose CAF intake was associated with increased frontoparietal network activation and FC in different working memory tasks (Koppelstaetter et al., 2008; Haller et al., 2013). It is important to note that the relationship between CAF and BOLD activity is not trivial because CAF acts not only as an excitatory neurostimulant but also as a vasoconstrictor reducing cerebral blood flow (Fredholm et al., 1999). This may explain why several studies also find CAF-induced widespread decreases in FC (Wong et al., 2012; Tal et al., 2013; Rack-Gomer et al., 2009).

However, up to date there are no studies investigating the cognitive enhancement effect of different stimulants in neither memory nor FC. To fill this gap, we used resting state fMRI and memory related behavioral data from a double blind randomized placebo-controlled study (Repantis et al., 2021). In this study, 48 male participants were randomized into three groups receiving a stimulant (CAF, MOD, MPH) or a placebo first in two separate sessions. Next to fMRI data acquisition, the participants performed a series of cognitive tasks including different memory tasks (see, Methods section and Repantis et al., 2021).

Given prior evidence, we assumed enhanced performance in memory tests as a function of all three stimulants. Due to the reported widespread effects of MPH, MOD and CAF on FC, we chose an agnostic, data-driven approach employing whole brain FC analyses on resting state fMRI data. However, since all three stimulants have repeatedly shown to affect prefrontal areas, we assumed that stimulant-induced changes in large-scale neural networks should involve prefrontal cortices. Additionally, stimulant-induced FC changes specific to memory should involve medial temporal lobe structures including the hippocampus, due to its consistent association with successful memory formation (Ranganath et al., 2005; for a meta-analysis, see Grady, 2020; Wais, 2008).

## Methods

### Participants

The study included 48 healthy, right-handed male participants (*M*=26.27, *SD*=3.47, range: 21-36). The participants were recruited via internet recruitment and screened for presence of psychiatric or medical disorders as determined by a physician and using the Mini-International Neuropsychiatric Interview (Sheehan et al., 1998). Only male participants were included due to prior evidence showing an interaction of the female hormone cycle with resting state FC as well as performance in memory tasks (Lisofsky et al., 2015). Further exclusion criteria were: regular or excessive consumption of caffeinated drinks (regularly drinking > 4 cups per day) within the last 6 months^1^, current usage of any medications, lifetime consumption of prescription stimulants (modafinil, methylphenidate) or illicit *hard* substances (e.g. cocaine, crack, heroine), consumption of other illicit substances (e.g. speed, amphetamine, THC, ecstasy, MDMA) within the past year, regular or irregular heavy smoking within the last 5 years [two subjects reported to smoke irregularly and little] as well as current irregular day-night rhythm (e.g. shift-work).

To further control for irregular sleeping patterns, bad sleep quality as well as strong differences in circadian rhythm between the participants, we administered the German translated versions of the Pittsburgh Sleep quality index (PSQI, Buysse et al., 1989) and the Morning-Eveningness Questionnaires (D-MEQ, Horne & Östberg, 1976; Griefahn et al., 2001). According to the PSQI, none of the participants self-reported an unhealthy sleeping pattern (*M*=2.46, *SD* = 1.65; max=5; note, subjects with values between 0-5 are considered healthy sleepers). Furthermore, according to the D-MEQ, the participants exhibited as similar circadian rhythm (*M*=2.52, *SD* = 0.46; max=3.47, min=1.63; note, the D-MEQ ranges from 0=*clearly “evening person”* to 6=*clearly “morning person”*).

Written informed consent was obtained from all subjects and they were financially compensated for their time of participation. The study was approved by the Berlin State Ethics Committee (LAGeSo Berlin, Germany; 13/0138-EK12) and conducted according to the Declaration of Helsinki (1964).

### Study design and procedure

We used a placebo-controlled, randomized, double-blind within-subject study design with three arms in which a stimulant was tested against placebo. Each participant received only one stimulant (and placebo) in the form of a white capsule for oral ingestion and was tested at two different occasions at the same time of the day in the early afternoon. The sessions were separated by about one week. The stimulants that were used were immediate-release MPH (20 mg), MOD (200 mg) and CAF (200 mg). The doses where chosen based on known equipotency in clinical practice and prior trials as well as (for MPH and MPD) similar occupancy of the dopamine transporter (Franke et al. 2017; Repantis et al. 2010; Theunissen et al. 2009; Volkow et al. 2009; Volkow et al. 1998).

The white capsules containing one of the stimulants or the placebo (=microcrystalline cellulose) had been prepared by a pharmacy of the Charité (Berlin) in such a manner that they looked identical and weighed the same. Participants encoded the memory task with visual material in the MRI scanner and completed the rest of the tasks (including retrieval of this memory task) afterwards outside of the scanner (see section *behavioral measures of memory*). The start of the fMRI procedure was 90 min after oral substance ingestion. This timeframe was chosen so that all three stimulants had reached approximately their peak concentration in blood during testing. Two resting state scans were acquired directly before and after the encoding of the memory task with visual material in the MRI scanner, hence, 90 min and 120 min after oral substance ingestion. The reason why we acquired two instead of one resting state scan was to account for general within-subject resting state variability (Bijsterbosch et al., 2017) and potential within-subject variation in elimination half-live of the stimulants (Wagner, 1973).

### Behavioral measures of memory

In the original study two different memory tasks were administered – memory of visual and audio material (see Repantis et al., 2021). For better comparability, we used the same memory outcome measures as reported in Repantis and colleagues (2021).

#### Memory task of visual material

In a declarative memory task, participants were exposed to 72 common nouns. The encoding difficulty was matched between lists and pre-tested. Based on previous research, word lists counting about 70 words prevent ceiling effects (Riedel and Blokland 2015). The words were presented in 12 blocks (six words each) inside the scanner. Each word was presented for 2000 ms with a jittered inter stimulus interval of 2 – 5 s. Blocks were interspersed with fixation periods. An early recall was performed outside the scanner approximately 20 min after learning phase.

#### Memory task of auditory material

Implicit and explicit verbal memory was measured with a false memory test outside of the scanner (Roediger and McDermott 1995). Participants heard five sets of 15 words each (75 words in total). Every set contained semantically similar words that could be associated with one critical lure which was not presented itself. As an example, the presented words “drive”, “street”, “key”, “garage”, etc. are all associated to the lure “car”. After the presentation an early recall was performed outside the scanner. Afterwards, participants underwent a recognition test in which a list of 40 words was presented. These included 20 words from the auditory material word list and 20 new words with the lures included among them. A sensitivity index d’ was calculated using the formula d’ = Z(hit rate) – Z(false alarm rate) in order to examine how well participants discriminated between old and new words. Hit rate was defined as (hits/(hits + misses) (hits were the correctly identified old words; misses were the old words that were not recognized)(Macmillan and Creelman 1990). False alarm rate was defined as (false alarms/(false alarms + correct negative) (false alarms were new words that were falsely identified as old words; correct negative were new words that were correctly identified as new words). For the Z transformation the formula NORM.S.INV(hit) – NORM.S.INV(false alarm) was used. Perfect scores were adjusted using the formula 1 – 1/(2n) for perfect hits and 1/(2n) for zero false alarms, with n being the number of total hits or false alarms (20 and 20 respectively) (Haatveit et al. 2010; Macmillan and Creelman 1990). A higher d’ score indicates that the signal was recognized better.

Retention of information was assessed for both memory tasks with visual and auditory material using a delayed free recall test for the two sets of learned words. This was conducted by telephone 24 hours after the session. Participants were informed about the upcoming late recall task but were asked not to actively try to retain the words.

### Behavioral data analysis

#### Analysis of single memory tasks

To analyze the stimulant-induced changes on memory, we first analyzed the outcome measures for each memory task separately in a General Linear Mixed Model (GLMM) [for a discussion of the advantages using the GLMM approach, see Barr, Scheepers & Tilly, 2013; Baayen, Davidson & Bates, 2008]. The memory measures which served as dependent variables for the single GLMMs were 1) d’ from the implicit memory task; 2-3) the amount of correctly remembered items in the memory task of visual material during early [early_visRecall] and late recall [late_visRecall]; 4-5) the amount of correctly remembered items in the memory task of audio material during early [early_audRecall] and late recall [late_audRecall] and 6-7) the amount of falsely remembered critical lures (false memories) in the memory task of audio material during early [early_false_audRecall] and late recall [late_false_audRecall]. D’ was modeled assuming a Gaussian error distribution. Except for d’, all other outcome measures [2-7] were count variables which is why we modeled those variables assuming a Poisson error distribution of the model residuals with the default log link function (Gardner et al., 1995).

The main independent variable of interest was the condition (binary: stimulant vs placebo). Other covariates of no interest included into the model were 1) the group (a categorical variable indicating the specific stimulant group MOD, MPH or CAF) and 2) the medication order (a categorical variable indicating in which order the participants received either the placebo or the stimulant first). Subjects were modeled as random intercepts.

To assess the influence of the condition (stimulant vs. placebo) on all seven memory outcome measures, we performed likelihood ratio tests of the full model with the condition variable against the baseline model without the condition variable. The baseline model included the group variable, order (stimulant or placebo in first session) and the random subject effect. The full model was identical to the baseline model with the exception of the additional condition variable. P-values for the individual models were obtained from likelihood ratio tests. Additionally, all obtained p-values from the likelihood ratio tests were corrected for multiple comparisons using the false discovery rate (Benjamini & Hochberg, 1995).

All statistical analyses were carried out in R (version 4.0.3, R Core Team, 2014). The GLMMs and linear models were estimated using the glmmTMB package (version 1.0.2.1; see Brooks et al., 2017). Visual inspection of residual plots did not reveal any obvious deviations from homoscedasticity or normality for all models. The behavioral data, the extracted within-connectivity values per cluster as well as the analysis code are publicly available on github https://github.com/MaxiBecker/Neuroenhancement.

#### Measurement model for latent memory change factor

To relate stimulant-induced changes in memory to changes in resting state FC, we calculated a latent memory change factor from the difference scores (stimulant – placebo) of the individual memory measures. The latent memory change factor represents the shared covariance of the individual memory tasks and is therefore a more robust measure for memory change as a function of the stimulants. We included the difference values of the following four memory measures into the final model: d’, early_audRecall, late_audRecall, early_visRecall (see section *Analysis of single memory tasks*). To reach a good model fit, we excluded the difference values of the following variables: late_visRecall, early_false_audRecall and late_false_audRecall and fixed the error variance of the variable early_audRecall to one (see Fig. 3).

The latent factor was estimated within a confirmatory factor analysis (CFA) in R using the lavaan package (version 0.6-7, Yves Rosseel, 2012). Model fit was evaluated via multiple fit indices: the χ^2^ goodness-of-fit statistic, Bentler’s comparative fit index (CFI), root mean square error of approximation (RMSEA) and standardized root-mean-square residual (SRMR).Accepted thresholds indicating good model fit are RMSEA <= .05, SRMR <0.1 and CFI >= .95 (Schermelleh-Engel et al., 2014; Hu and Bentler, 1998, 1999).

### Rs-fMRI parameters

Imaging was performed on a Siemens 3T Magnetom Trio Scanner (Siemens Healthcare, Erlangen, Germany) using an echo planar protocol with a 12-channel radiofrequency head coil. Functional resting state images were obtained using a T2*-weighted echo planar imaging (EPI) sequence sensitive to blood oxygen level dependent (BOLD) contrast (TR = 2000 ms, TE = 30 ms, slice thickness = 3mm, image matrix = 64 × 64, FOV =216 mm, flip angle = 80°, voxel size = 3mm^3^, 36 axial slices). The participants were instructed to relax and keep their eyes closedfor both resting state scans. Each scan contained a total of 150 volumes.

Structural images were collected using a three-dimensional T1-weighted magnetization prepared gradient-echo sequence (MPRAGE) (repetition time (TR) = 2500 ms; echo time (TE) = 4.77 ms; slice thickness = 1mm, acquisition matrix = 256 × 256 × 192, flip angle = 7°; FOV = 256 mm, voxel size = 1mm^3^).

### Rs-fMRI preprocessing and denoising

Rs-fMRI state data was preprocessed, denoised and subsequently analyzed using CONN – an open source connectivity toolbox (version 19c, Whitfield-Gabrieli & Nieto-Castanon, 2012).

#### Preprocessing

The data was processed using CONN’s default preprocessing pipeline which includes the following steps: Functional scans were realigned and slice-time corrected. Realignment includes co-registering all scans and resampling them to the first reference image using b-spline interpolation (see Anderson et al., 2001). Outlier scans were identified using framewise displacement and CONN’s default parameters (framewise displacement above 0.9mm or global BOLD signal changes above 5 standard deviations).

Functional and structural data was normalized into MNI space and segmented into gray matter, white matter as well as CSF tissue. Finally, the functional data was smoothed using an 8mm full width half maximum Gaussian kernel to increase BOLD signal to noise-ratio.

#### Denoising

Confounding effects to the estimated BOLD signal were first estimated and subsequently removed for each voxel in each subject and for each condition as well as each session using Ordinary Least Squares regression. For this purpose, an anatomical component-based noise correction procedure (CompCor) as implemented in CONN was used. Potential confounding effects included: six estimated subject-motion parameters including their first-order derivatives, noise components from areas of cerebral white matter and cerebrospinal fluid, constant or linear session effects as well as the above-mentioned identified outlier scans. Note, no global signal regression was applied to avoid the risk of artificially introducing anticorrelations (negative connectivity) into the FC estimates (Chai et al., 2012). Furthermore, the effects of denoising were further manually evaluated using CONN’s quality control plots. Finally, the resulting time series were additionally band-pass filtered to .008 - .09 Hz.

### Rs-fMRI data analysis

#### First-level analysis in CONN

We adopted a whole brain data-driven approach because we did not have a specific hypothesis where in the brain FC should increase as a function of all three stimulants. To investigate those stimulant-induced changes in FC on a network level, i.e. the whole brain, we parcellated the brain into 268 Regions of Interest (ROIs) using the Shen atlas (Shen et al., 2013). The Shen atlas is parcellated based on resting state FC and therefore ideal for network analysis using FC (Finn et al., 2015; Rosenberg et al, 2016; Shen et al., 2013). For this reason, the Shen atlas was preferred to CONN’s default FSL Harvard-Oxford atlas that contains only 132 ROIs derived from structural data (106 cortical and subcortical areas and 26 cerebellar areas from the AAL Atlas) (Finn et al., 2015; Rosenberg et al, 2016; Shen et al., 2013).

The time series for each ROI were acquired by mean averaging the BOLD time series of each voxel that belongs to the respective ROI. ROI-to-ROI connectivity matrices (268×268) were computed for each participant (48), for each condition (drug vs. placebo), each scan (2) and each session (2) separately. Each element in each connectivity matrix represents the strength of FC between a pair of ROIs and was defined as the Fisher-transformed bivariate correlation coefficient between the preprocessed and denoised BOLD time series of two ROIs.

#### Second-level analysis in CONN

Rather than focusing on single connections between all possible pairs of ROIs, we adopted a hierarchical cluster analysis as implemented in CONN which categorizes ROIs into clusters based on ROI-to-ROI functional similarity and anatomical proximity metrics (Sørensen, 1948). This method is based on a complete-linkage metric meaning that the distance between two clusters is defined as the maximum distance between all pairs of ROIs within the two clusters. CONN’s default distance measure that we used is defined as D= 0.95*D_func_ + 0.05*D_anat_ where D_func_ is the squared Euclidean distance between the connectivity patterns for every pair of ROIs (averaged across all subjects), and D_anat_ represents the squared Euclidean distance between the anatomical centroid coordinates for every pair of ROI.

To infer stimulant-induced changes in FC on these groups/clusters of ROIs, a multivariate parametric General Linear Model was performed for all connections included in each of these groups/clusters of ROIs (in terms of within- and between network connectivity sets, see Jafri et al, 2008). The resulting F-statistic for each pair of related ROI-groups/clusters was thresholded at an FDR-corrected cluster level *p*<.05 (Benjamini and Hochberg, 1995). To characterize the pattern of individual connections, a post-hoc uncorrected *p*<.05 height threshold (connection-level) was applied.

To investigate stimulant induced changes on FC, we contrasted the stimulant condition (2 scans) with the placebo condition (2 scans) averaged for each stimulant group (between subject contrast: 0.33*MOD + 0.33*MPH + 0.33*CAF).

We additionally investigated whether there were whole-brain stimulant-induced changes in FC that relate to stimulant-induced changes in general memory. For this, we first extracted the subject-specific factor loadings of the latent memory change factor (see section *Latent Memory change factor*). Before entering the second level in CONN, the values were normalized, mean centered and orthogonalized to all three stimulant groups. Subsequently, the values were correlated with every group/cluster of related ROIs in the brain while controlling for differences between the groups (between subject contrast: 1*Memory + 0*MOD + 0*MPH + 0*CAF). Finally, those correlations were contrasted between the stimulant condition (2 scans) and the placebo condition (2 scans).

For exploratory purposes, the resulting (two) clusters from the group analysis in CONN were further examined to investigate to what extent they also relate to change in latent memory performance. To this end, we extracted the within-FC of each cluster (mean average of all ROI-pairs that belong to the respective cluster). For each cluster, we subtracted the resulting within-FC values of the stimulant condition from the placebo condition. Subsequently, the difference values of those clusters were entered into a separate linear model together with three covariates of no interest to predict change in latent memory performance (i.e., the individual factor loadings from latent memory change factor). Due to high collinearity (r=0.53, 95%CI [0.392 - 0.649]), the difference values of both clusters could not be entered as predictors into the same linear model. Therefore, we performed two separate linear models each predicting change in latent memory performance. The additional covariates per model were a binary variable indicating the respective scan and the order in which the subjects received either the stimulant or the placebo (binary variable: stimulant first vs placebo first). P-values for the individual models were obtained from likelihood ratio tests. That is to say, a baseline model including only the two covariates of no interest (scan and order [stimulant or placebo in first session]) was compared to the full model 1 including the two covariates and the difference value of the first cluster. The baseline model was also compared to the full model 2 including the two covariates and the difference value of the second cluster. To control for multiple comparison, all obtained p-values from the likelihood ratio tests were corrected using the false discovery rate (Benjamini & Hochberg, 1995).

## Results

### Effects of stimulants on memory

Details on the model estimates from the individual GLMMs are depicted in tables S1-S3 in the supplementary material.

#### Memory task with visual material

On average, participants correctly recalled 35.21 (*SD*=18.24) words in the stimulant and 31.69 (*SD*=16.88) words in the placebo condition. This difference in early memory recall was statistically significant [*Chi*^*2*^ (1) = 8.622, *p-FDR*=.008].

After 24 hours participants correctly recalled 21.53 (*SD*=17.61) words in the stimulant and 16.77 (*SD*=13.33) words in the placebo condition on average. This difference in late memory recall was also significant [*Chi*^*2*^ (1) = 30.87, *p-FDR*= .000] (see Fig. 1).

**Figure 1.**
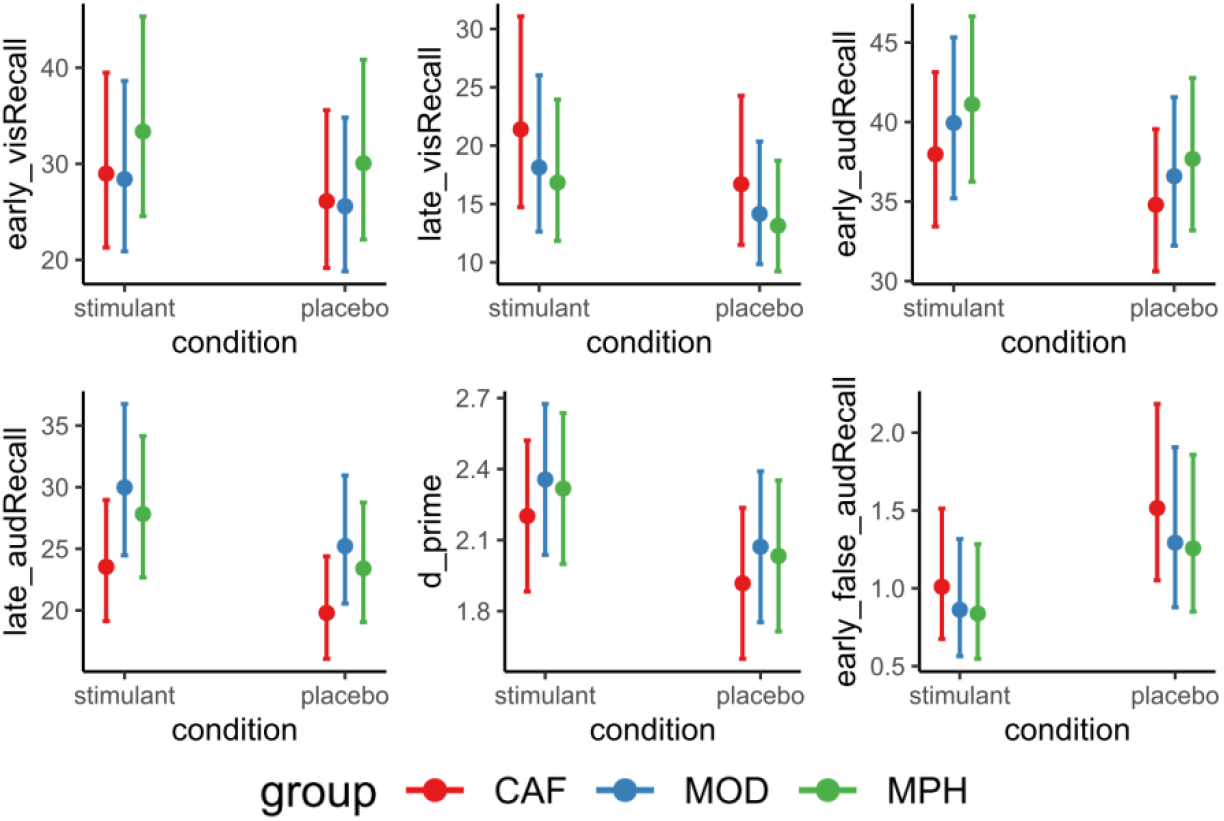
Marginal means of memory performance in memory tasks with visual and audio material as a function of condition plotted separately for all three stimulant groups. *Note*. CAF = caffeine, MOD = modafinil, MPH = methylphenidate; error bars represent between-subject 95% confidence intervals. D_prime = d’ from an implicit memory task; early_audRcall = early recall for memory task with audio material; late_audRecall = late recall for memory task with audio material; early_visRecall = early recall for memory task with visual material; early_false_audRecall = early recall for lures (false memory) in memory task with audio material.

#### Memory task with audio material

On average, participants correctly recalled 40.60 (*SD*=9.64) words in the stimulant and 37.21 (*SD*=10.21) words in the placebo condition. This difference in early memory recall was statistically significant [*Chi*^*2*^ (1) = 7.116, *p-FDR*=.013].

After 24 hours participants correctly recalled 28.96 (*SD*=11.97) words in the stimulant and 24.35 (*SD*=10.48) words in the placebo condition on average. This difference in late memory recall was significant [Chi^2^ (1) = 19.11, *p-FDR*=.000] (see Fig. 1).

#### Memory task with audio material– false memory

Furthermore, on average participants falsely remembered 0.91 (*SD*=0.89) lure words in the stimulant condition and 1.38 (*SD*=1.18) lure words in the placebo condition. This difference in early false memory recall was statistically significant [Chi^2^(1) = 4.43, *p-FDR*=.041] (see Fig. 1).

After 24 hours, participants falsely remembered 1.18 (*SD*=1.05) lure words in the stimulant and 1.44 (*SD*=1.11) lure words in the placebo condition. However, this difference in late false memory recall was not statistically significant [Chi^2^ (1) = 2.17, *p-FDR*=.141].

#### Implicit memory task

On average, d’ in the stimulant condition was 2.29 (*SD*=0.76) whereas d’ was 2.01 (*SD*=.71) in the placebo condition. This difference was statistically significant [*Chi*^*2*^ (1) = 5.880, *p-FDR*=.015] (see Fig. 1).

#### Measurement model for latent memory change factor

The model converged normally after 31 iterations. All variables loaded in the same direction (positive). Except for *early_visRecall*, all standardized factor loadings were within moderate to high range (difference values for *d’:* λ=.479; *early_audRecall:* λ =.992; *late_audRecall:* λ =.567; *early_visRecall:* λ =.181), suggesting that those variables contributed significantly to the latent memory change factor (see Fig. 3, left panel). The exact test statistic did not reach significance [χ^2^ = 0.588, *p* = .899], indicating that the model was not significantly different from the data. Additionally, practical fit indices [CFI = 1.0, RSME = < .001 and SRMR = .027] also suggested a good fit of the model to our data.

### Effects of stimulants on functional connectivity

The hierarchical cluster analysis revealed two significant clusters of ROIs that were all positively correlated. The first cluster [*F*(2,44) = 11.31, *p-FDR* = .0237] consisted of 26 connections from predominantly medial parietal regions being functionally connected to right prefrontal regions as a function of all three stimulants combined (see Fig. 2, cluster 1). A post-hoc analysis of the condition and group specific effect sizes revealed that those parietal-prefrontal regions were negatively correlated in the placebo condition, while this negative correlation was significantly reduced in the stimulant condition (see Fig. 2, cluster 2). The second cluster [*F*(2,44) = 12.25, *p-FDR* = .0237] consisted of 10 connections from predominantly lateral parietal regions that were positively functionally connected to right prefrontal regions (see Fig. 2). A post-hoc analysis of the condition and group specific effect sizes revealed that FC generally increased as a function of all three stimulants (see Fig. 2). Table S5 in the supplementary material lists all ROIs belonging to each cluster including their individual T-statistics. To demonstrate that the results from the hierarchical cluster analysis (as shown in Fig. 2) were not just driven by one stimulant, we additionally plotted both clusters separately for each group, each condition and each scan in Figure S4 in the supplementary material.

**Figure 2.**
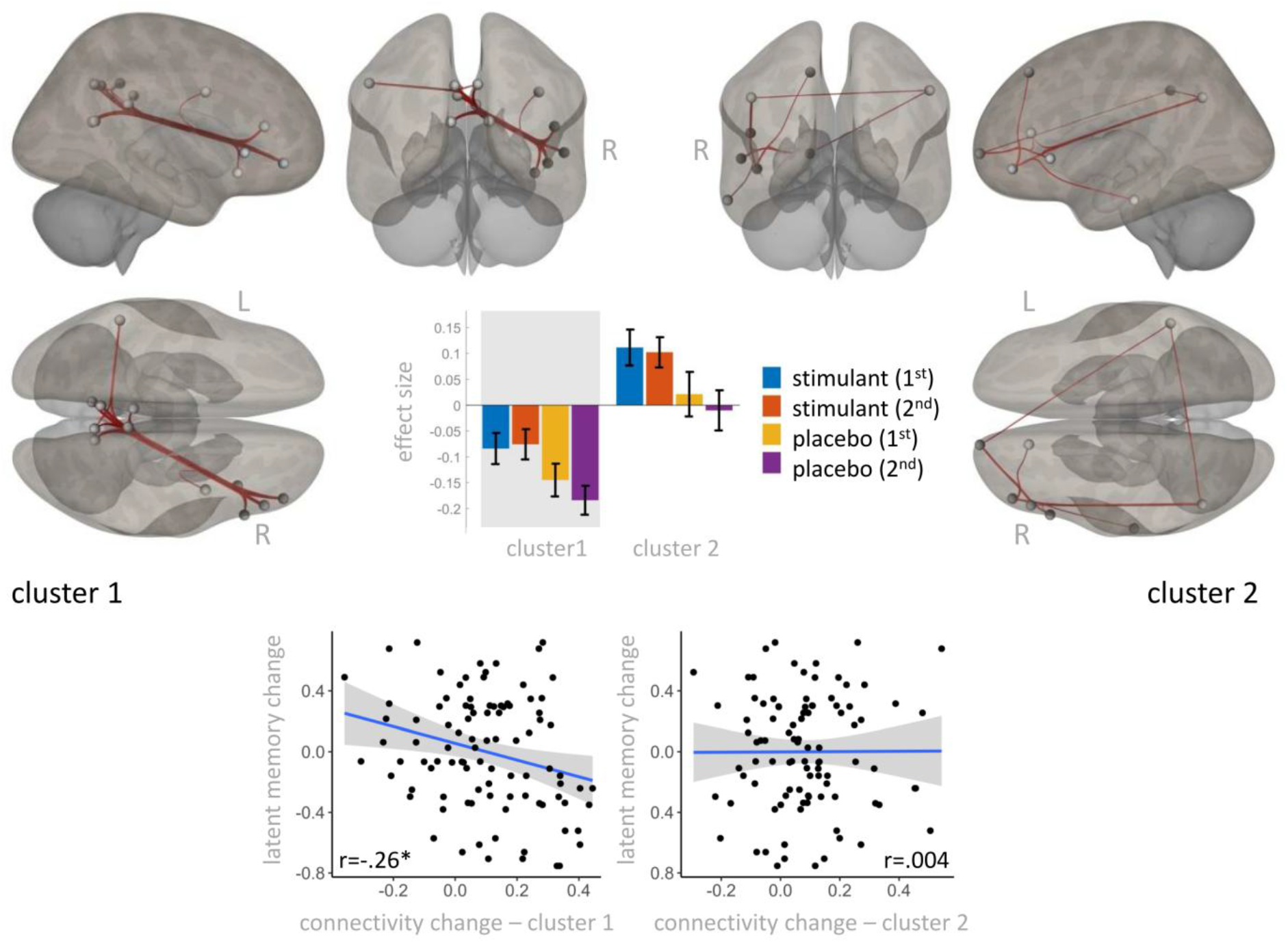
Effects of caffeine, modafinil & methylphenidate on functional connectivity. *Note*. Upper panel: Results from hierarchical cluster analysis. Two clusters significantly differed between the stimulant and placebo condition. 1^st^ and 2^nd^ refer to the first and second resting state scan. The red lines represent significant correlations of the time series between the regions of interest (gray spheres). Lower panel: Change in functional connectivity from the placebo to the stimulant condition correlated significantly negatively with change in latent memory performance from the placebo to the stimulant condition in cluster 1 but not in cluster 2. * indicates significance at a p-level <.05 (FDR-corrected).

For exploratory purposes, we investigated whether stimulant-induced change in FC in both clusters predicts the amount of latent memory change. The first cluster [*Chi*^2^(1) =7.33, *p-FDR*= 0.016] negatively predicts latent memory change. The negative relationship is reasonable given that the ROI pairs in this cluster in the placebo condition are more negatively connected than in the stimulant condition (see Fig. 2, lower left panel). In contrast, FC change in the second cluster [*Chi*^2^(1)= 0.07, *p-FDR*=0.799] does not predict latent memory change (see Fig. 2, lower right panel). Details on the model estimates from the linear model are presented in Table S4 in the supplementary material. For the sake of completeness, the relationship between latent memory change and the respective cluster plotted separately for each group is presented in Figure S5 in the supplementary material.

### Link between stimulant-induced changes in whole brain rs-FC and memory

When investigating which brain areas correlate more strongly with stimulant-induced changes in latent memory performance in the stimulant compared to the placebo condition, one cluster survived multiple comparisons on a whole brain level [*F*(2,43) = 11.75, *p-FDR* = .037]. This cluster consisted of 16 connections between ROIs in visual (lingual, calcarine, middle occipital cortex) and medial temporal lobe visual areas (Hippocampus, Parahippocampus and Superior Temporal Pole) (see Fig. 3). A post-hoc analysis revealed that ROIs from both areas were negatively correlated in the placebo condition, while this negative correlation was significantly reduced in the stimulant condition (see Fig. 3). Table S5 in the supplementary material lists all ROIs belonging to each cluster including their individual T-statistics.

**Figure 3.**
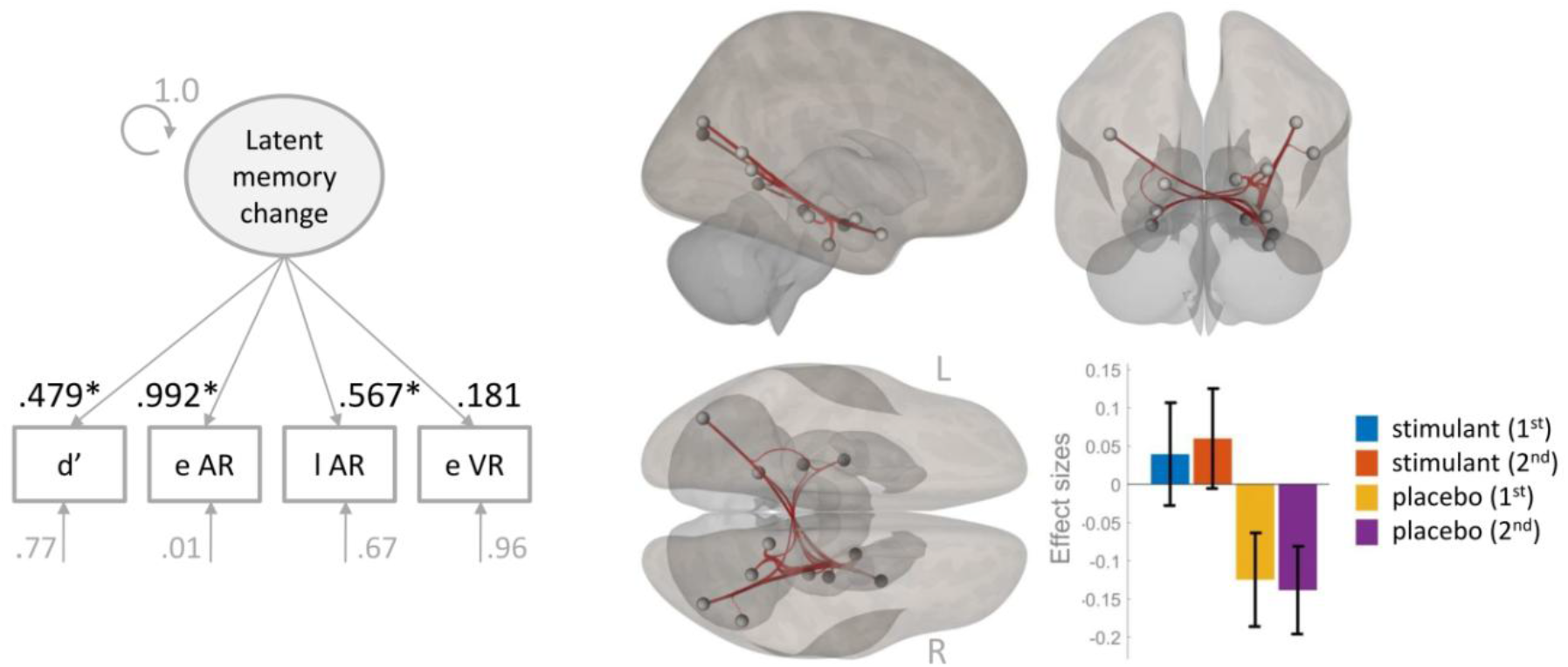
Correlation of stimulant-induced changes in whole-brain rs-FC with changes in latent memory. *Note*. Left panel: Measurement model for latent memory change factor. d’ = d prime from implicit memory task; e AR = early recall for memory task with audio material; l AR = late recall for memory task with audio material; e VR = early recall for memory task with visual material; * signifies statistical significance at the *p*<.05 level. Right panel: Results from whole brain hierarchical cluster analysis. Negative correlation of occipital – medial temporal lobe cluster with the latent memory change factor is significantly reduced in the stimulant compared to the placebo condition. 1^st^ and 2^nd^ refer to the first and second resting state scan, respectively. The red lines represent significant correlations of the time series between the regions of interest (gray spheres).

## Discussion

In this study, we investigated stimulant-induced effects of MPH, MOD and CAF on memory and related FC. Despite their different modes of action, all three stimulants are considered cognitive neuroenhancers due to their positive effects on attention, cognitive control and specifically memory (Frank & Lieb, 2010; for a critical review on this matter, see Repantis et al., 2010). However, mnemonic and brain-functional effects of cognitive neuroenhancers (e.g. MPH, MOD and CAF) are usually studied separately. While this expands our knowledge of stimulant-specific effects on behavior and brain function, the question remains, whether there are shared cognitive enhancement effects over and above the individual stimulant. We speculated that one cognitive enhancement mechanism could be the stimulants’ direct or indirect impact on extracellular levels of catecholamines in the brain. Specifically, dopamine has been shown to shape large-scale neural networks including prefrontal regions underlying cognitive functions. For this reason, we utilized behavioral and resting state fMRI data from a double-blind randomized placebo-controlled study (see Repantis et al., 2021) and investigated the combined effects of MPH, MOD and CAF on memory and FC at rest (Repantis et al., 2021). We hypothesized that memory performance should increase and large-scale neural networks including prefrontal regions should be modulated as a function of all three stimulants. Additionally, we assumed that stimulant-induced FC changes specific to memory should involve medial temporal lobe structures due to their consistent association with memory formation (for a meta-analysis, see Grady, 2020).

As predicted, we found enhanced mnemonic performance in different memory tasks in the stimulant compared to the placebo condition. That is to say, implicit memory measured as d’ was enhanced. Correct recall for visual and audio material was also enhanced in the stimulant condition immediately after encoding as well as 24 hours later. Furthermore, false memory for lures was reduced in the stimulant-condition, but only after immediate not during late recall. Further congruent with our hypotheses, we found a stimulant-induced effect of increased FC. Hierarchical cluster analysis revealed two main clusters of connections between predominantly prefrontal and parietal ROIs indicating FC change between the fronto-parietal network (FPN) and default mode network (DMN) at rest (see Fig. 2 and Table S5). In one cluster, FC between predominantly lateral parietal and right prefrontal ROIs was increased in the stimulant compared to the placebo condition. In the other cluster, negative FC between predominantly medial parietal (Precuneus, Posterior Cingulum (PCC)) DMN and right prefrontal FPN ROIs was decreased in the stimulant compared to the placebo condition. A post-hoc analysis revealed that the amount of negative connectivity reduction from the stimulant to the placebo condition predicted the amount of performance change in latent memory. On a whole brain level, however, one specific cluster including connections between medial temporal lobe areas (e.g. Hippocampus and Parahippocampus) also belonging to the DMN and visual areas was significantly related to stimulant-induced latent memory change. That is to say, in this cluster negative FC was significantly reduced in the stimulant compared to the placebo condition.

### Stimulant-induced changes in coupling between FPN and DMN

The DMN is a set of cortical regions comprising hubs mostly located in the ventromedial prefrontal, inferior parietal lobule and PCC/Precuneus (Greicius et al., 2003). Those nodes exhibit coherent fluctuations during resting state and have been associated with a range of cognitive processes including self-oriented mental activity and autobiographical as well as episodic memory retrieval (Buckner et al., 2008; Sestieri et al., 2011). Prior studies report that this network can be modulated by different stimulants such as MPH (Liddle et al., 2011; Silverstein et al., 2016) as well as MOD (Minzenberg et al., 2011; Schmaal et al., 2013) and CAF (Wong et al., 2012; Wu et al., 2014).

The FPN comprises regions such as the anterior insular cortex, anterior prefrontal, dorsolateral prefrontal and dorsomedial superior frontal cortex/anterior cingulate and the anterior inferior parietal lobule (Dang, O’Neil & Jagust, 2012). It has been primarily associated with attention and cognitive control (Dodds, Morein-Zamir & Robbins 2010; Scolari, Seidl-Rathkopf & Kastner 2015; Badre and D’Esposito, 2007; Cabeza et al., 2008). However, Iidaka and colleagues have shown evidence that this network is also associated with successful retrieval in episodic memory (Iidaka et al., 2006). FC within the FPN has been shown to be modulated by different stimulants such as MOD (Visser et al., 2019; Esposito et al., 2012); CAF (Koppelstaetter et al., 2008; Haller et al., 2013) and MPH (Mehta et al., 2000; Müller et al., 2014). In fact, Schmidt and colleagues report a broad recruitment of frontoparietal regions when MOD and MPH are consumed compared to placebo (Schmidt et al., 2017).

At rest, the DMN typically exhibits anticorrelated activity with the FPN as we have observed in cluster 1 in the placebo condition (Chai et al., 2012; Fox et al., 2005; Uddin et al., 2009). Importantly however, the FPN has been found to flexibly couple with the DMN, depending on the task domain supporting goal-directed cognitive processes (Spreng et al., 2010). Dopaminergic signal transmission may function as one underlying mechanism in this regard. Evidence suggests that dopamine enhances coupling between the FPN and DMN at rest (Dang et al., 2012). This finding is in line with our assumption that one neurobiological denominator of the cognitive enhancement effect could lie in the stimulants’ ability to increase extracellular levels of catecholamines (e.g. dopamine, noradrenaline) in the brain. Catecholaminergic innervations range across the brain with high overlapping concentrations in hippocampus and prefrontal cortex (for review see Ranjbar-Slamloo & Fazlali, 2020). Hence, it may be the ability of specifically FPN’s right prefrontal hubs to flexibly couple and decouple from the DMN depending on the task demands that relate to the cognitive enhancement effects associated with all three stimulants (CAF, MOD, MPH).

Consequently, we found that the amount of negative connectivity reduction between predominantly right prefrontal DMN and medial posterior FPN regions (Precuneus, PCC) significantly predicted stimulant-induced latent memory change. While the right IFG is implicated in attention to facilitate goal-directed behavior (Dodds, Morein-Zamir & Robbins 2011), parietal regions of the DMN support episodic memory retrieval (Sestieri et al., 2011). Hence, decreasing the anticorrelation (negative connectivity) between both regions may be associated with increased attention during successful encoding of the stimulus material, leading to enhanced memory performance in the stimulant condition. However, the functional associations coupling between the nodes of the FPN and DMN seem to be specific because we only found a latent memory change related association between medial (not lateral) parietal DMN and right prefrontal FPN regions. This is congruent with studies, showing that the DMN is not a functionally homogenous unity but that different nodes within this network contribute distinctively to cognitive processes like cognitive control (Leech et al., 2011; Laird et al., 2009)

Further evidence, supporting the assumed relationship between catecholamines especially dopamine, FPN-DMN coupling and memory comes from patients with attention-deficit/hyperactivity-disorder (ADHD). ADHD has been associated with weakened prefrontal cortex function (Arnsten, 2006), altered DMN-FPN interactions (Sripada, Kessler & Angstadt, 2014) and deficient attention-related encoding and retrieval processes (Ortega et al., 2020). Importantly, ADHD is successfully treated with MPH by increasing the extracellular concentration of catecholamines in prefrontal, striatal and other brain areas (Mehta et al., 2000).

### Memory related stimulant-induced FC changes between visual and medial temporal lobe regions

Episodic memory is critically dependent on the MTL and its functional connections to the cortex. For example, anatomical studies find many direct connections between the Parahippocampus (PHC) and the DMN (Burwell, 2000; Witter et al., 2000). The MTL is also strongly functionally connected to early and late visual areas (Wang et al., 2016; Cordova, Tomary & Turk-Browne, 2016).

Cordova and colleagues (2016) show that memory retrieval can be facilitated by attention-related increased coupling between visual areas and MTL regions during encoding of visual stimuli. In line with those results, we find stimulant-induced modulations in FC between MTL and visual areas to be directly related to memory enhancement. Those stimulant-induced modulations may also be associated with attention, however, we did not observe increased positive but decreased negative connectivity between visual and MTL regions. While positive connectivity between brain regions at rest is assumed to reflect synchronized activity between those regions, the physiological mechanisms underlying negative connectivity are still debated (Goelman, Gordon, Bonne, 2014). Next to non-neuronal hemodynamic processes, one physiological source for negative connectivity could be neuronal inhibition (Bianciardi et al., 2011; Shmuel et al, 2006; Devor et al, 2007). Hence, the decreased negative connectivity between visual and MTL regions in the stimulant condition could reflect reduced inhibition between those brain regions facilitating encoding of the stimulus material in both memory tasks. In both tasks, participants had to memorize visually or aurally presented words. In fact, the Lingual Gyrus – one visual region identified within the hierarchical cluster – plays an important role in the recognition of words (Michelli et al., 2000) and has also been found to support encoding of words via visual imagery (Leshikar et al., 2012). However, further research is necessary to investigate negative connectivity between MTL and visual regions in the context of memory.

### Limitations

It is worth mentioning three study limitations that may reduce the generalizability and interpretability of the results. First, only male participants were recruited to avoid differential menstrual cycle effects on the two study sessions 7 days apart. This selectivity also reduces study-irrelevant within-subject variability regarding the rs-fMRI data (Lisofsky et al., 2015). However, research including women is necessary to test whether the stimulant-induced behavioral and FC results generalize to the whole population. Similarly, we only included participants with no to low/irregular caffeine consumption into the study to avoid cognitive and physiological side effects due to caffeine withdrawal during the time period of the experiment (see James & Rogers, 2005; James, 2014). However, about 90% of the general U.S. population report to regularly consume caffeine (Meredith et al., 2013).

Second, the sample size per stimulant group (n=16) was rather low, thus complicating any interpretation of differential effects of any of the three stimulant. We did not find evidence for systematic differences between the groups in the sense that one stimulant group was driving the obtained overall results on memory and FC. However, given that the combined cognitive enhancement effect of all three stimulant groups only explained about 4-10% of the variance in memory, it cannot be excluded that there are systematic differences between the stimulant groups if the group size was increased (e.g. n= 100).

Third, we only used self-reports to screen the participants for confounding variables such as caffeine, prescription stimulant or illicit substance consumption or sleep quality. More objective measures such as sleep diaries or actigraphy to evaluate participants’ sleeping patterns as well as a urine drug screen prior to testing would have been a better control for those confounding variables.

## Conclusion

We investigated cognitive enhancement effects of stimulants (CAF, MPH, MOD) on memory and related large-scale neural networks. Our preliminary results indicate that reduced negative FC between the FPN and DMN as well as between visual and MTL regions reflect a cognitive enhancement effect on the neural system-level specific to stimulant-induced memory enhancement. Further research is required to test, whether those enhanced memory-related changes in FC are generalizable to other pharmacological neuroenhancers. Finally, further research is required to investigate the shared stimulant-induced effects on different neurotransmitter systems such as catecholamines (dopamine) that give rise to such large-scale neural-network changes.

## Data Availability

The behavioral data, the extracted within-connectivity values per cluster as well as the analysis code are publicly available on github https://github.com/MaxiBecker/Neuroenhancement.

## Acknowledgements

This work was supported by a grant (No. 85648) of the Volkswagen Foundation, Germany. The Volkswagen Foundation had no role in the design, data collection, data analysis, data interpretation, or writing of the manuscript. S.K. has been funded by the German Science Foundation (DFG SFB 936/C7), the European Research Council (ERC-2016-StG-Self-Control-677804) and a Fellowship from the Jacobs Foundation (JRF 2016-2018). M.B. was supported by the Einstein Foundation Berlin (EPP-2017-423). We acknowledge support by the Open Access Publication Fund of the Humboldt-University Berlin.

We want to thank Lukas C. Adam for help in collecting the data.

The authors declare no conflict of interest.

## Supplementary material

**Table S1.** Influence of Stimulant condition on correct early and late recall - visual material.

|  | early recall - visual material |  |  | late recall - visual material |  |  |
| --- | --- | --- | --- | --- | --- | --- |
| Predictors | IRR | CI | p | IRR | CI | p |
| (Intercept) | 21.91 | 15.52 – 30.93 | <0.001 | 15.50 | 9.96 – 24.11 | <0.001 |
| group [MOD] | 0.98 | 0.65 – 1.49 | 0.926 | 0.85 | 0.51 – 1.42 | 0.542 |
| group [MPH] | 1.18 | 0.78 – 1.78 | 0.444 | 0.80 | 0.48 – 1.33 | 0.391 |
| **condition [stimulant]** | **1.11** | **1.04 – 1.19** | **0.003** | **1.28** | **1.17 – 1.41** | **<0.001** |
| med_order [plc 1st] | 1.42 | 1.01 – 1.99 | 0.043 | 1.14 | 0.75 – 1.73 | 0.534 |
| Random Effects |  |  |  |  |  |  |
| $\sigma^2$ | 0.03 | | | 0.20 | | |
| ICC | 0.91 |  |  | 0.69 |  |  |
| Marg.R<sup>2</sup> / Cond. R<sup>2</sup> | 0.092 / 0.916 |  |  | 0.043 / 0.707 |  |  |

*Note.* IRR = Incidence Rate Ratio; Est. = estimates; med\_order [plc 1st] = medication order [placebo was administered in the first session]; $\sigma^2$ = variance; ICC = intraclass coefficient; CI= 95% confidence interval, p-values are uncorrected for multiple comparison.

**Table S2.** Influence of Stimulant condition on early and late recall - audio material.

|  | early recall |  |  | late recall |  |  |
| --- | --- | --- | --- | --- | --- | --- |
| Predictors | IRR | CI | p | IRR | CI | p |
| (Intercept) | 34.00 | 29.37 – 39.35 | <0.001 | 18.94 | 14.90 – 24.08 | <0.001 |
| group [MOD] | 1.05 | 0.88 – 1.25 | 0.566 | 1.28 | 0.96 – 1.70 | 0.095 |
| group [MPH] | 1.09 | 0.91 – 1.29 | 0.351 | 1.19 | 0.89 – 1.58 | 0.237 |
| **condition [stimulant]** | **1.09** | **1.02 – 1.16** | **0.008** | **1.19** | **1.10 – 1.29** | **<0.001** |
| med_order [plc 1st] | 1.05 | 0.91 – 1.21 | 0.520 | 1.09 | 0.86 – 1.38 | 0.466 |
| Random Effects |  |  |  |  |  |  |
| $\sigma^2$ | 0.03 | | | 0.04 | | |
| ICC | 0.65 |  |  | 0.79 |  |  |
| Marg.R<sup>2</sup> / Cond. R<sup>2</sup> | 0.045 / 0.669 |  |  | 0.096 / 0.809 |  |  |

*Note.* IRR = Incidence Rate Ratio; med\_order [plc 1st] = medication order [placebo was administered in the first session]; $\sigma^2$ = variance; ICC = intraclass coefficient; CI= 95% confidence interval; p-values are uncorrected for multiple comparison.

**Table S3.** Influence of Stimulant condition on implicit memory (d’) and early false memory (lures)

| Predictors | implicit memory - d' |  |  | early false recall (lures) |  |  |
| --- | --- | --- | --- | --- | --- | --- |
|  | Est. | CI | p | IRR | CI | p |
| (Intercept) | 1.84 | 1.48 – 2.20 | <0.001 | 1.79 | 1.23 – 2.61 | 0.003 |
| group [MOD] | 0.15 | -0.27 – 0.57 | 0.471 | 0.85 | 0.54 – 1.34 | 0.492 |
| group [MPH] | 0.13 | -0.29 – 0.55 | 0.560 | 0.81 | 0.52 – 1.28 | 0.372 |
| **condition [stimulant]** | **0.28** | **0.06 – 0.51** | **0.012** | **0.67** | **0.46 – 0.98** | **0.037** |
| med_order [plc 1st] | 0.15 | -0.20 – 0.49 | 0.405 | 0.72 | 0.49 – 1.05 | 0.091 |
| Random Effects |  |  |  |  |  |  |
| $\sigma^2$ | 0.31 | | | 0.64 | | |
| ICC | 0.41 |  |  | 0.00 |  |  |
| Marg.R<sup>2</sup> / Cond. R<sup>2</sup> | 0.055 / 0.438 |  |  | 0.104 / NA |  |  |

*Note.* Est. = Estimates; IRR = Incidence Rate Ratio; med\_order [plc 1st] = medication order [placebo was administered in the first session]; $\sigma^2$ = variance; ICC = intraclass coefficient; CI= 95% confidence interval; p-values are uncorrected for multiple comparison.

**Table S4.** Relationship between stimulant-induced functional connectivity change in both clusters and stimulant-induced change in latent memory performance.

| Predictors | latent memory change |  |  | latent memory change |  |  |
| --- | --- | --- | --- | --- | --- | --- |
|  | Est. | CI | p | Est. | CI | p |
| (Intercept) | -0.12 | -0.24 – 0.00 | 0.054 | -0.17 | -0.29 – -0.05 | 0.006 |
| **fcon change - cluster 1** | **-0.51** | **-0.89 – -0.14** | **0.008** |  |  |  |
| **fcon change - cluster 2** |  |  |  | **0.06** | **-0.38 – 0.49** | **0.799** |
| scan [2] | 0.01 | -0.12 – 0.14 | 0.864 | -0.00 | -0.14 – 0.14 | 0.970 |
| med_order [Plc 1st] | 0.34 | 0.21 – 0.47 | <0.001 | 0.35 | 0.21 – 0.49 | <0.001 |
| R<sup>2</sup> / R<sup>2</sup> adjusted | 0.273 / 0.249 |  |  | 0.215 / 0.190 |  |  |

*Note.* Est. = Estimates; CI= 95% confidence interval; p-values are uncorrected for multiple comparison. med\_order [plc 1st] = medication order [placebo was administered in the first session].

**Figure S4.**
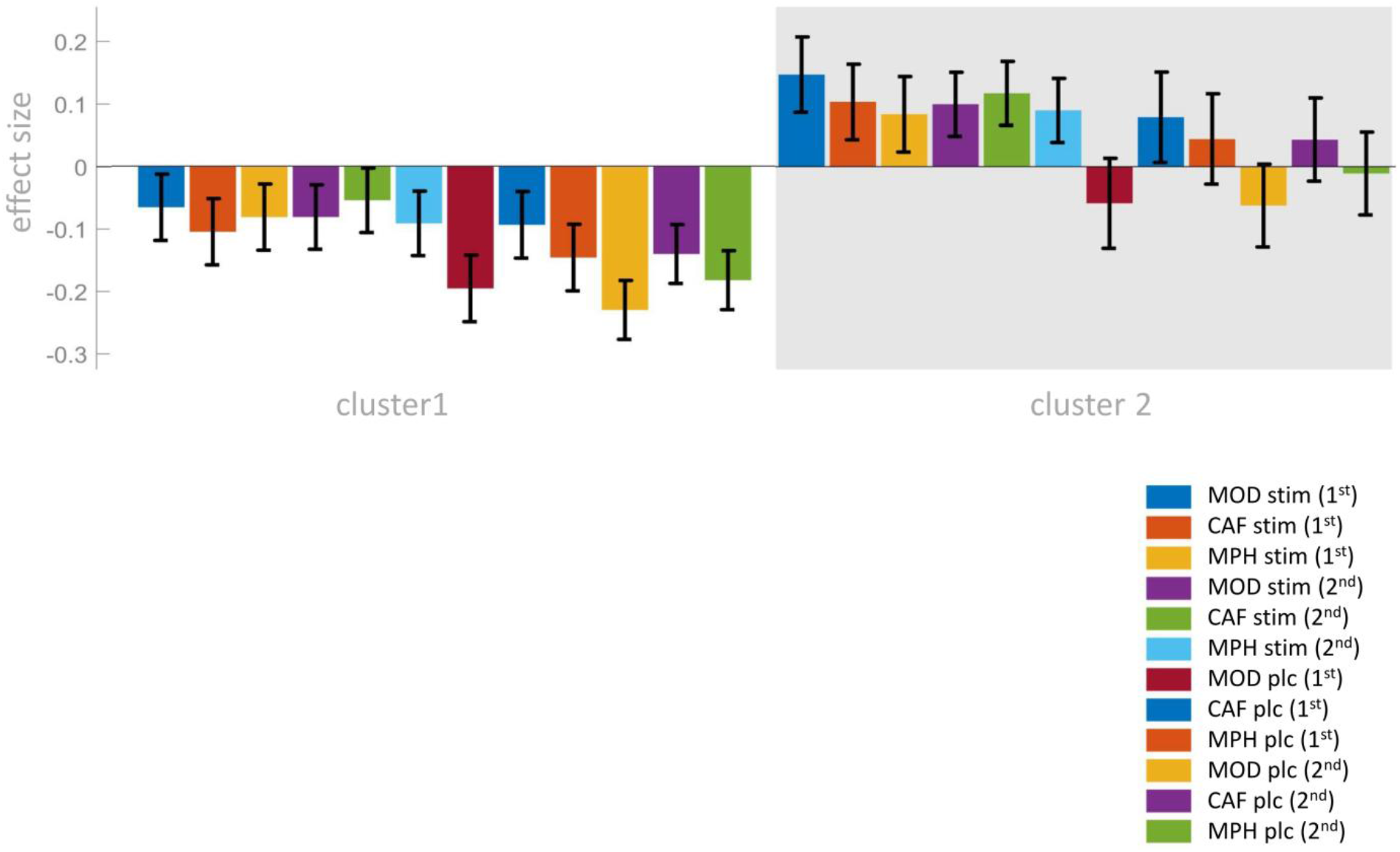
Effects of caffeine, modafinil & methylphenidate on functional connectivity– plotted separately for all three stimulants. *Note*. Results from whole brain hierarchical cluster analysis. CAF = caffeinegroup, MOD = modafinil group, MPH = methylphenidate group. stim = condition in which respective stimulant was administered; plc = condition in which placebo was administered; 1^st^ and 2^nd^ refer to the first and second resting state scan, respectively.

**Figure S5.**
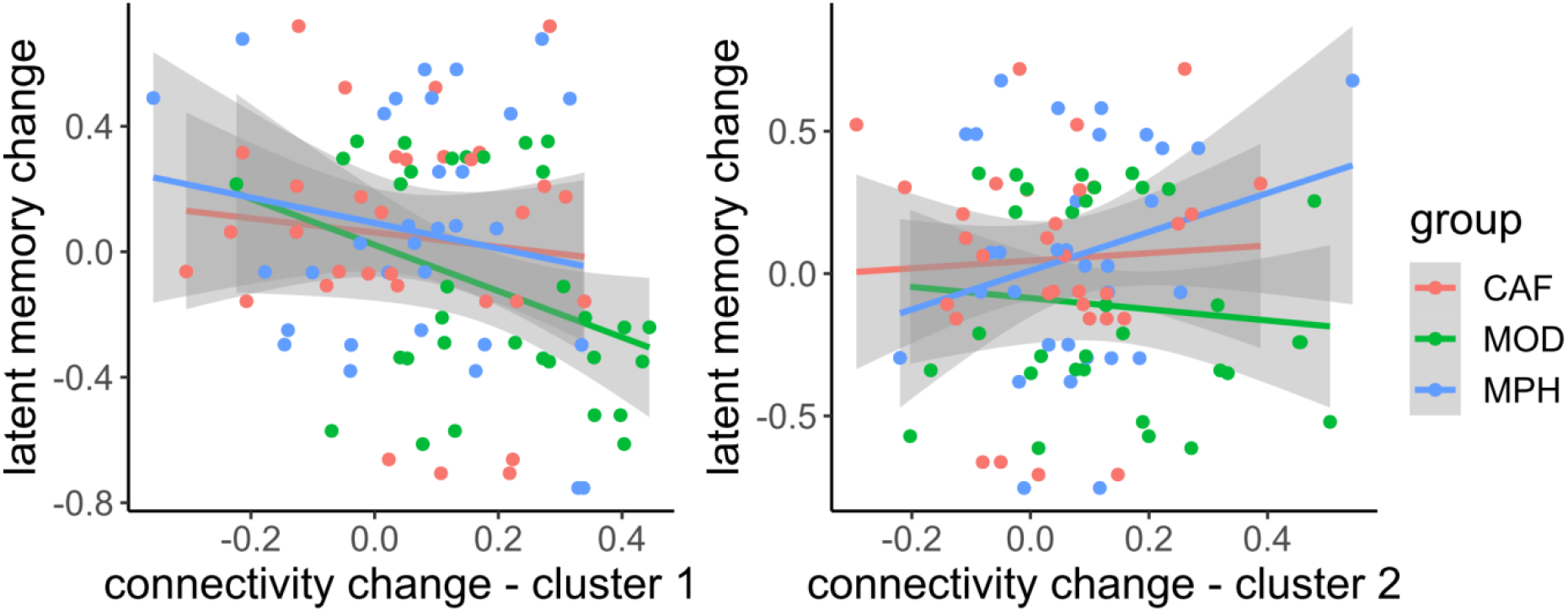
Relationship between both FC clusters from hierarchical cluster analysis and latent memory change plotted separately for all three stimulants. *Note*. CAF = caffeine, MOD = modafinil, MPH = methylphenidate.

**Table S5.**
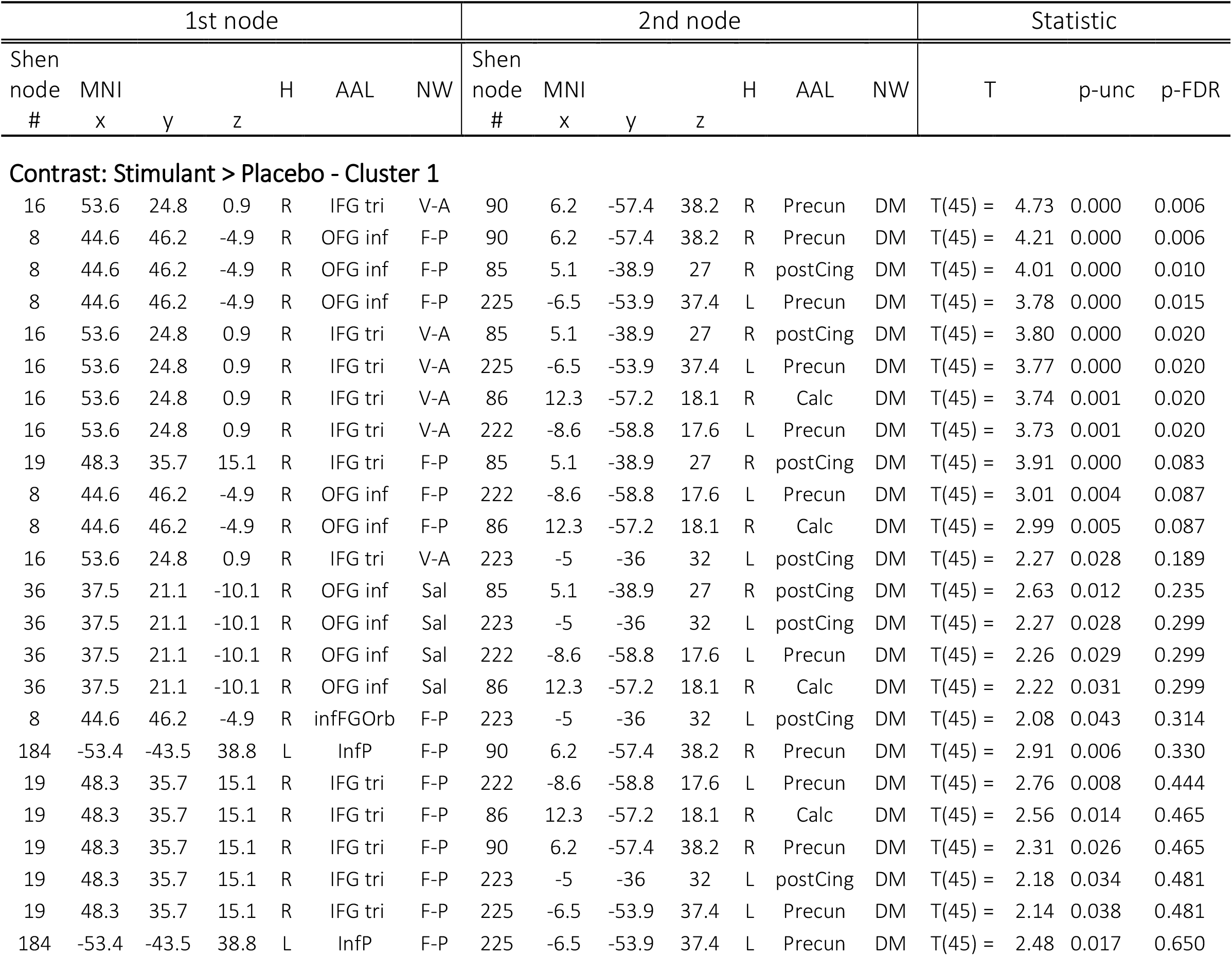
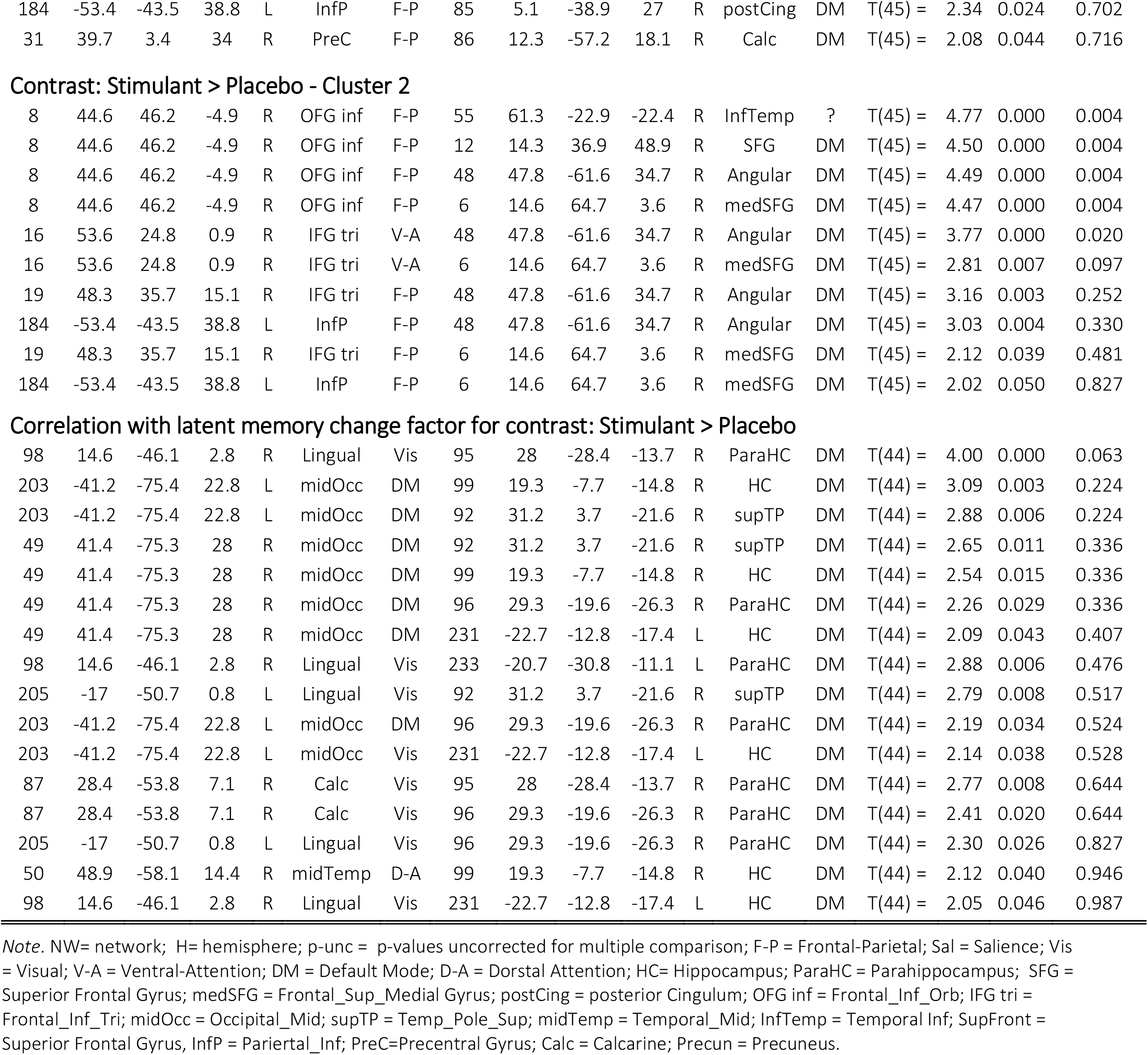
Individual connections between cluster nodes from hierarchical cluster analysis.

## Footnotes

1 Participants were instructed to refrain from drinking any caffeinated drinks throughout the entire length of the study. However, this affected only three participants who reported to irregularly consume few caffeinated drinks (45 participants reported not to consume any caffeinated drinks currently at all).

